# Coenrollment of Critically Ill Patients in PROSPECT: A Protocol and Statistical Analysis Plan

**DOI:** 10.1101/2025.03.29.25324889

**Authors:** Alex Thabane, Diane Heels-Ansdell, Nicole Zytaruk, Deborah Cook, the PROSPECT Investigators, the Canadian Critical Care Trials Group

## Abstract

**Introduction:** The enrollment of a patient into more than one study (i.e., coenrollment) has risks which warrant exploration, particularly with respect to possible effects on trial outcomes. This pre-planned secondary analysis will examine the sensitivity of treatment effects to coenrollment in an international critical care trial (Probiotics: Prevention of Severe Pneumonia and Endotracheal Colonization Trial (PROSPECT).

**Objective(s):** The primary objective is to evaluate the sensitivity of the effect of probiotics on the primary endpoint of VAP to patient coenrollment in at least one other study. The secondary objectives are to describe the characteristics of coenrolled patients and the studies they were coenrolled into; examine any differences in baseline traits; understand differences in center-level characteristics between coenrolling and non-coenrolling centers; identify factors associated with coenrollment; and explore the relationship between coenrollment status and the incidence of adverse events.

**Methods:** We developed a protocol and statistical analysis plan (SAP) for this secondary analysis involving the conduct of a Cox regression model, including treatment allocation, coenrollment status, and the interaction between the two as independent variables. We also describe our planned statistical analyses for the secondary objectives, involving descriptive statistics, univariable analyses, and multivariable analyses.

**Ethics and Dissemination:** The results of this study will be published in a peer-reviewed journal focused on critical care research or trial methodology, and presented at local, national, and international conferences. As a secondary analysis, this study does not require research ethics board approval. All data will be presented in aggregate and without patient identifiers.

## Introduction

Probiotics have been suggested as a cost-effective strategy for preventing ventilator-associated pneumonia (VAP) in critically ill patients [1], with several systematic reviews of randomized controlled trials reporting evidence of efficacy [2-4]. However the Probiotics: Prevention of Severe Pneumonia and Endotracheal Colonization Trial (PROSPECT) [5], which included 2,650 critically ill patients randomized to receive either the probiotic *Lactobacillus rhamnosus GG* or a placebo, found no effect of probiotics on the development of ventilator-associated pneumonia (VAP). This finding has called into question the utility of probiotics for the prevention of VAP.

Given the incongruence between the PROSPECT findings and prior randomized controlled trials, it is important to assess the robustness of the primary analyses in PROSPECT through sensitivity analyses [6]. One variable which could have modified the results is coenrollment, defined as the simultaneous or sequential enrollment of one patient into more than one study. While coenrollment confers benefits in terms of research efficiency, several concerns have been raised regarding the coenrollment of patients into multiple studies, including the interaction of study drugs and the subsequent impact of those interactions on treatment effect estimates [7-9]. Exploring the sensitivity of the treatment effect of probiotics to coenrollment will help strengthen the robustness of the conclusion that probiotics have no clinical value in the prevention of VAP.

We present a protocol and statistical analysis plan for secondary analysis of the PROSPECT dataset with the objective of examining the sensitivity of the effect of probiotics on the primary outcome of ventilator-associated pneumonia (VAP) to coenrollment.

## Methods

### Study Design

This study is a pre-planned secondary analysis of PROSPECT, a multi-center, international, randomized placebo-controlled trial [5].

### Objectives

#### Primary Objective

The primary objective is to evaluate the sensitivity of the effect of probiotics on the primary endpoint of VAP to patient coenrollment.

#### Secondary Objectives

The secondary objectives are to describe the characteristics of coenrolled patients and the studies they were coenrolled into; explore differences in baseline patient characteristics between coenrolled and non-coenrolled patients; explore differences in center-level characteristics between coenrolling and non-coenrolling centers; identify patient- and center-level factors associated with patient coenrollment; and explore the relationship between coenrollment and adverse events.

### Summary of PROSPECT Trial

#### Eligibility Criteria

Patients included in PROSPECT met the following inclusion criteria, as outlined in the trial protocol and statistical analysis plan (SAP) [10]:

1. Adults ≥ 18 years of age admitted to a medical, surgical, or trauma intensive care unit (ICU).
2. Receiving invasive mechanical ventilation, estimated to be required for ≥72 hours.

Patients were excluded from participation for meeting any one of the following criteria:

1. Invasive mechanically ventilated >72 hours at the time of screening.
2. Potential increased risk of iatrogenic probiotic infection including specific immunocompromised groups: HIV < 200CD4 cells/µL, chronic immunosuppressive medications, previous transplantation at any time, chemotherapy in the last 3 months, absolute neutrophil count < 500 cells/µL.
3. Risk for endovascular infection: rheumatic heart disease, congenital valve disease, surgically repaired congenital heart disease, unrepaired cyanotic congenital heart disease, valvular replacement (mechanical or bio-prosthetic), previous or current endocarditis, permanent endovascular devices (e.g., endovascular grafts, inferior vena cava filters, dialysis vascular grafts), tunnelled hemodialysis catheters, pacemakers, or defibrillators. The following were not exclusions: coronary artery stents or bypass grafts, mitral valve prolapse, bicuspid aortic valve, temporary catheters (central venous, peripherally inserted, extra-corporeal life support-related) or neurovascular coils.
4. Primary diagnosis of severe acute pancreatitis.
5. Percutaneously inserted feeding tubes in situ.
6. Strict contraindication or inability to receive enteral medications.
7. Intent to withdraw advanced life support.
8. Previous enrolment in this trial or current enrollment in a potentially confounding trial.

#### Recruitment & Randomization

Patients were recruited from 44 intensive care units ICUs in Canada, the United States, and Saudi Arabia. Informed consent was obtained from each patient or their substitute decision-maker. Patients were randomly allocated in a 1:1 ratio via a computer-based random number generator to receive either the study intervention (i.e., the probiotic *Lactobacillus rhamnosus GG*) or an enteral placebo [10]. Randomization was stratified by center and by admission category (i.e., medical, surgical, trauma) [10].

#### Interventions

Patients in the probiotics group received 1 × 10^10^ colony forming units of *Lactobacillus rhamnosus GG* (i-Health, Inc.) in one capsule suspended in tap or sterile water, administered through a nasogastric or orogastric feeding tube [10]. Patients in the placebo group were randomized to receive an identical capsule containing microcrystalline cellulose, delivered the same way [10]. Patients received their allocated treatment for 60 days in the ICU, or until ICU discharge, death, or isolation of *Lactobacillus* spp. in a culture from a sterile site or if the sole or predominant organism in a culture from a non-sterile site [10].

#### Primary Endpoint

The primary endpoint of PROSPECT was VAP, defined as the presence of new, progressive, or persistent radiographic infiltrate on chest radiographs after at least 2 days of mechanical ventilation, plus at least 2 of the following criteria: a fever above 38°C; white blood cell count less than 3 × 10^6^ or exceeding 10 × 10^10^; or purulent sputum [5].

### Blinding

Patients, bedside clinicians, investigators, and research coordinators were blinded to the treatment allocation during this trial. While the statistician involved in the primary analysis of the trial was blinded to the treatment allocation until the main analysis was complete, for this pre-planned secondary analysis study, the statistical team will be unblinded to the treatment allocation.

### Statistical Analysis

To meet our primary objective, we plan to conduct a statistical analysis similar to that performed in the primary trial – a Cox regression model adjusted for center and admission diagnosis, with an independent variable of treatment and dependent variable of VAP [5] – but including as additional independent variables patient coenrollment status and the interaction between treatment allocation and coenrollment status. This analysis will allow us to statistically test the sensitivity of the effect of probiotics to coenrollment. We will report the hazard ratios (HRs) with corresponding 95% confidence intervals (CIs) between coenrolled and non-coenrolled patients in both treatment groups and report the interaction p-value between treatment allocation and coenrollment status. We hypothesize that coenrollment will not influence the treatment effect of probiotics on the primary endpoint of VAP.

As a secondary objective, we plan to describe the characteristics of coenrolled patients, including the number of coenrolled patients and number of studies in which patients were coenrolled, by treatment allocation and overall. We also plan to describe the characteristics of coenrolled studies (i.e., studies into which PROSPECT patients were coenrolled), including the informed consent model of these studies, study design, study affiliation, and funding source. Descriptive statistics will be reported as counts (percentages) and means (standard deviations [SDs]) for categorical and continuous variables, respectively.

We plan descriptive statistics to describe the differences in baseline patient characteristics between coenrolled and non-coenrolled patients. Patient-level variables of interest include: age; APACHE II score [11]; Clinical Frailty Score [12]; inotrope or vasopressor infusion; dialysis use; individual granting informed consent. We similarly have planned descriptive statistics to explore differences in center-level characteristics between coenrolling and non-coenrolling centers. Center-level variables of interest include: center size (i.e., number of ICU beds); years of PROSPECT participation; year of PROSPECT initiation; site investigator trial experience; lead research coordinator trial experience; hospital type; and location.

To identify factors associated with patient coenrollment, we plan to conduct a multi-level logistic regression analysis, with a random intercept for center to account for clustering at the center-level. The dependent variable will be coenrollment status. We will include the following center-level variables as independent variables: center size, years of PROSPECT participation, site investigator trial experience, lead research coordinator trial experience, and hospital type (academic vs university). We will also include the following patient-level variables: age, sex, APACHE II score, and individual granting informed consent (substitute decision-maker vs. patient). We will report odds ratios (ORs) with corresponding 95% CIs and p-values. We hypothesize that coenrollment will be positively associated with less seriously ill patients (as measured by the APACHE II score), larger center size, informed consent granted via substitute decision-maker, academic centers, and centers with high site investigator and lead research coordinator experience. We hypothesize that no association will be found between years in which the center participated in PROSPECT, patient age, or sex.

To explore the association between coenrollment status and adverse events, we will report the rate of adverse by coenrollment status (counts with percentages) and conduct a Fisher’s exact test comparing the rate of adverse events between coenrolled and non-coenrolled patients, reporting p-values. We hypothesize that coenrollment will have no association with adverse events.

The analysis will be performed by the lead author (AT), to be replicated independently by the trial analyst (DH) who will be blinded to the results. A summary of the statistical analysis plan is presented in **Appendix A1**.

### Timeline

This study is a secondary analyses of data from a large, international randomized trial [5]. While participant recruitment, data collection, and results for the primary trial have been reported, the conduct of this secondary analyses has yet to be completed. We anticipate that this study, requiring data analyses and manuscript development, will be completed and submitted for publication in mid-2025.

## Ethics & Dissemination

### Ethics and Safety Considerations

Being a secondary analysis of an RCT database requiring no additional information and analyzing only anonymized data, this study did not undergo secondary research ethics board (REB) approval. The initial trial was approved by Health Canada, the REBs of all participating hospitals, and Public Health Ontario [10]. All data will be presented in aggregate and without any patient identifiers to ensure the anonymity of patients.

### Dissemination Plan

We plan to publish the results of this study in a peer-reviewed journal focused on critical care or trial methodology. The first author (AT) and co-primary investigator for the trial (DC) will lead knowledge translation efforts, mainly through the presentation of the results at local and national conferences. Efforts will be made to discuss the findings and their implications with relevant stakeholders (e.g., critical care trialists, patients, substitute decision-makers).

## Discussion

While the issue of coenrollment is relatively unexplored in the field of critical care, we have previously examined patterns, predictors, and consequences of coenrollment in two international critical trials: the OSCILLation for ARDS Treated Early Trial (OSCILLATE) and the PROphylaxis for ThromboEmbolism in Critical Care Trial (PROTECT) [8, 13]. In neither trial did coenrollment influence any of the safety or efficacy outcomes [8, 13]. However, multivariable analyses of factors associated with coenrollment yielded some differences in results between the two studies. In the ARDS trial, age, site investigator and research coordinator trial experience, center size, and country were associated with coenrollment in multivariable analyses [8]; in the thromboprophylaxis trial, coenrollment was not associated with age, but was associated with disease severity, center size, individual granting informed consent, center affiliation with a research consortium, and research coordinator experience [13]. While differences in model design (i.e., the selection of independent variables included in the model) may explain some of these discordant findings, this can be taken as evidence of the complexity of coenrollment decision-making and conditional context. Exploration of factors associated with coenrollment in each trial is therefore warranted.

Coenrollment is handled variably by ICUs and institutions. Policies against the coenrollment of patients have been previously reported [14], suggesting hesitation due to possible negative effects. Beyond treatment interactions, increased patient burden continues to be a key apprehension when considering coenrollment [14]. Understanding the effects of coenrollment in trials such as PROSPECT, and the factors associated with coenrollment, will help critical care trialists better understand this practice, including the types of patients unlikely to be coenrolled, and may help investigators provide patients and research sites with accurate information on the possible benefits and harms of coenrollment.

This study has several strengths. PROSPECT was unique in capturing detailed coenrollment data, including the number of coenrollment events and characteristics of coenrolled studies, which is uncommon among critical care trials. PROSPECT also collected factors associated with coenrollment in other studies, such as investigator and trial coordinator expertise [13], which permits exploring whether these factors are associated with coenrollment in PROSPECT. Further, our proposed multi-level logistic regression analysis will allow evaluation of the independent association of several patient- and center-level factors with coenrollment whilst controlling for correlation in observations within study sites. Controlling for within-site correlation has the potential to improve the validity of the model, subsequently leading to more accurate estimations of association. This study could offer a prototype analysis to explore coenrollment in future randomized trials.

## Conclusion

This study will generate evidence about the consequences, patterns, and predictors of coenrollment in a large critical care trial, contributing to the wider discussion on the patterns, harms, and benefits of coenrollment.

## Data Availability

No data is presented in this manuscript.

## Supporting Information

**S1 File. Statistical Analysis Table**

## Acknowledgements

We are grateful to the patients and families participating in this trial, as well as the collaborating Research Coordinators and Investigators, and bedside clinicians who supported this work. The trial was designed by the PROSPECT Steering Committee, the PROSPECT Investigators and Research Coordinators and the Canadian Critical Care Trials Group. We would like to thank the Methods Center staff for their expertise with PROSPECT data management, including Nicole Zytaruk, Lois Saunders, Shelley Anderson-White, Alyson Takaoka, Mary Copland, France Clarke, Lori Hand, Megan Davis, Neala Hoad, Melissa Shears and Kristine Wachmann.

