## Appendix A1 for "Coenrollment of Critically Ill Patients in PROSPECT: A Protocol and Statistical Analysis Plan"

**Appendix A1: Summary of Statistical Analysis Plan**

| **Objectives** | **Outcome** | **Explanatory Variables** | **Hypothesis** | **Methods/Reporting** |
| --- | --- | --- | --- | --- |
| **Primary** | | | | |
| 1) Evaluate the sensitivity of the effects of probiotics to patient coenrollment in the primary outcome | Ventilator Associated Pneumonia (VAP) | Treatment; Coenrollment Status | Co-enrollment will not influence the treatment effect of probiotics on the outcome of VAP | Cox Regression   - Adjusted for center and admission diagnosis. - Reporting HRs with 95% CIs - Reporting interaction p-value between treatment allocation and coenrollment status |
| **Secondary** | | | | |
| 1) Describe characteristics of coenrolled patients | N/A | Number of Coenrolled Patients; Number of Studies in Which Patients Were Coenrolled | N/A | Descriptive Statistics   - Counts (with percentages) - Means (with standard deviations) |
| 2) Describe characteristics of coenrolled studies | N/A | Informed Consent Model; Design; Study Affiliation; Funding Source | N/A | Descriptive Statistics   - Counts (with percentages) - Means (with standard deviations) |
| 3) Explore differences between coenrolled and non-coenrolled patients | Patient Coenrollment | Age; sex; APACHE II score; clinical frailty score; inotrope or vasopressor infusion; dialysis use; informed consent by substitute decision-maker | N/A | Descriptive Statistics   - Counts (with percentages) - Means (with standard deviations) |
| 4) Explore differences between coenrolling and non-coenrolling centers | Center Coenrollment | Center Size; Years of PROSPECT Participation; Year of PROSPECT initiation; Site Investigator & Lead Research Coordinator Trial Experience; Hospital Type; Location | N/A | Descriptive Statistics   - Counts (with percentages) - Means (with standard deviations) |
| 5) Identify factors associated with coenrollment | Patient Coenrollment | Center-Level   - Center Size (# of ICU Beds); Years of PROSPECT Participation; Site Investigator and Lead Research Coordinator Experience; Hospital Type   Patient-Level   - Age; Sex; APACHE II Score; Informed Consent Grantor | Coenrollment will be associated with:   1) Less seriously ill patients (APACHE II) 2) Consent granted by an SDM 3) Academic centers 4) Larger center size 5) Experienced site investigators and lead research coordinators   Coenrollment will not be associated with:  1) Age  2) Sex  3) Years of PROSPECT participation | Multi-level logistic regression   - Random intercept for center - Reporting ORs with 95% CIs and p-values |
| 6) Explore relationship between coenrollment and adverse events | Adverse Events | Coenrollment Status | Coenrollment will not be associated with adverse events | Descriptive statistics  Counts (with percentages) by group and coenrollment status  Fisher’s Exact test   - Reporting p-values |
| *VAP = Ventilator-Associated Pneumonia; SDM = substitute decision-maker; ICU = intensive care unit; PROSPECT = Probiotics: Prevention of Severe Pneumonia and Endotracheal Colonization Trial; APACHE II = Acute Physiology And Chronic Health Evaluation II Score; OR = Odds Ratio; HR = Hazard Ratio; CI = confidence interval* | | | | |
